# Seropositivity to Nucleoprotein to detect SARS-CoV-2 infections: a tool to detect breakthrough infections after COVID-19 vaccination

**DOI:** 10.1101/2021.10.05.21264555

**Authors:** Lotus L. van den Hoogen, Gaby Smits, Cheyenne C.E. van Hagen, Denise Wong, Eric R.A. Vos, Michiel van Boven, Hester E. de Melker, Jeffrey van Vliet, Marjan Kuijer, Linde Woudstra, Alienke J. Wijmenga-Monsuur, Corine H. GeurtsvanKessel, Susanne P. Stoof, Daphne Reukers, Lisa A. Wijsman, Adam Meijer, Chantal B.E.M. Reusken, Nynke Y. Rots, Fiona R.M. van der Klis, Robert S. van Binnendijk, Gerco den Hartog

## Abstract

**Background:** With COVID-19 vaccine roll-out ongoing in many countries globally, monitoring of breakthrough infections is of great importance. Antibodies persist in the blood after a severe acute respiratory syndrome coronavirus 2 (SARS-CoV-2) infection. Since COVID-19 vaccines induce immune response to the Spike protein of the virus, which is the main serosurveillance target to date, alternative targets should be explored to distinguish infection from vaccination.

**Methods:** Multiplex immunoassay data from 1,513 SARS-CoV-2 RT-qPCR-tested individuals (352 positive and 1,161 negative) with a primary infection and no vaccination history were used to determine the accuracy of Nucleoprotein-specific immunoglobulin G (IgG) in detecting past SARS-CoV-2 infection. We also described Spike S1 and Nucleoprotein-specific IgG responses in 230 COVID-19 vaccinated individuals (Pfizer/BioNTech).

**Results:** The sensitivity of Nucleoprotein seropositivity was 85% (95% confidence interval: 80-90%) for mild COVID-19 in the first two months following symptom onset. Sensitivity was lower in asymptomatic individuals (67%, 50-81%). Participants who had experienced a SARS-CoV-2 infection up to 11 months preceding vaccination, as assessed by Spike S1 seropositivity or RT-qPCR, produced 2.7-fold higher median levels of IgG to Spike S1 ≥14 days after the first dose as compared to those unexposed to SARS-CoV-2 at ≥7 days after the second dose (p=0.011). Nucleoprotein-specific IgG concentrations were not affected by vaccination in naïve participants.

**Conclusions:** Serological responses to Nucleoprotein may prove helpful in identifying SARS-CoV-2 infections after vaccination. Furthermore, it can help interpret IgG to Spike S1 after COVID-19 vaccination as particularly high responses shortly after vaccination could be explained by prior exposure history.

## Introduction

Since late 2020, multiple countries have initiated vaccine roll-out against COVID-19 which is caused by severe acute respiratory syndrome coronavirus 2 (SARS-CoV-2) [1]. Breakthrough infections have been reported shortly after completion of the vaccination regimen [2]. Although COVID-19 vaccines were developed to prevent severe disease and mortality and not to provide sterile protection, it will remain important to monitor the frequency of breakthrough infections as well as their transmission potential, specifically as new SARS-CoV-2 variants emerge [3]. During the acute phase of an infection, molecular (RT-qPCR) and antigen tests are used to confirm symptomatic and asymptomatic breakthrough infections, i.e. after contact tracing or travel to a high-risk area. However, asymptomatic persons who do not seek testing will likely be missed. To ensure a complete picture of the frequency of breakthrough infections for surveillance purposes, frequent RT-qPCR testing would be needed which is time- and labor intensive as well as burdensome to individuals.

Serological assays can identify specific antibodies which indicate previous infection with SARS-CoV-2. SARS-CoV-2 serostatus can be determined high-throughput with multiplex immunoassays (MIA [4]), irrespective of the presence of clinical symptoms. Immunoglobulin G (IgG) antibodies persist for months after infection which widens the window of detection as compared to RT-qPCR and antigen tests [5]. This should provide a more accurate estimate of ongoing transmission in the general population. However, since the main serological marker used to date for SARS-CoV-2 is also the vaccine target, Spike S1 or RBD, alternative serological targets should be explored to distinguish past infection from vaccination. Nucleoprotein is one of the structural immunogenic SARS-CoV-2 proteins. Others have reported sensitivity estimates ranging from 70% to 96% with specificity at ≥95%, depending the assay and reference population used [6-9]. Reference populations consisted of healthcare workers or hospitalized patients, which are not representative for the general population. Moreover, patients with severe symptoms produce higher antibody levels than those with mild or no symptoms which leads to overestimation of sensitivity estimates [5, 10]. Hence, the reliability of Nucleoprotein to detect mild or asymptomatic infections, which represent the majority of COVID-19 cases [11, 12], is still unknown.

We previously described a bead-based detection method for simultaneous IgG detection to Spike S1 and Nucleocapsid [4]. In this study we aimed to determine the accuracy of seropositivity to Nucleoprotein and Spike S1 by time since RT-qPCR-confirmed infection with mild or asymptomatic SARS-CoV-2 using a prospective household survey as well as a nationwide population survey.

## Methods

### Study design and population

#### Household cohort of infected and noninfected participants

A prospective cohort study was performed in households where one household member had tested positive for SARS-CoV-2 to determine within household transmission [13]. Patients with a RT-qPCR-confirmed SARS-CoV-2 infection (=index case) in the Municipal Health Service (GGD) Utrecht region, central Netherlands, were invited to participate with their household if they had at least one child under the age of 18 living at home. Households from 54 index cases were enrolled from March 24^th^ to May 24^th^ 2020 (with a total of 242 participants). Households were excluded if one or more of the household contacts did not want to participate in the study upfront. Furthermore, infants under the age of 1 were excluded. Most families were those of healthcare workers, for whom RT-qPCR testing was available at enrolment during the first pandemic wave (March/April 2020). Study nurses visited the families at their household within 24 hours after inclusion (T1), 2-3 weeks after inclusion (T2) and 4-6 weeks after inclusion (T3) to collect a venous blood sample for serological testing as well as a naso- and oropharyngeal swab, oral fluid and they supplied a feces collection kit. At 9-11 months after T1 (T5), another venous blood sample was collected at which point some of the participants had been COVID-19 vaccinated. Participants filled out a questionnaire at each sampling timepoint including data on demographic factors, symptoms and symptom onset, and vaccination data where applicable (brand product, number of vaccinations and their dates). The study was ethically approved by the Medical-Ethical Review Committee of the University Medical Center Utrecht (NL13529.041.06). All participants above the age of 12 gave written informed consent. Both parents or guardians of participating children below the age of 16 also gave written informed consent for participation of the child.

#### National cohort of noninfected, convalescent and vaccinated individuals

Serum samples were collected in an ongoing, nationwide longitudinal serosurveillance study; the PIENTER Corona (PICO) cohort study described by Vos et al. [14, 15]. Briefly, the PICO study emanated from a large-scale nationwide cross-sectional study performed in 2016-17 (PIENTER-3 [16]). Participants from the PIENTER-3 study who had consented to follow-up were invited to participate in the PICO study in April 2020 [14] and the cohort was extended with an additional nationwide random sample in June 2020 [15]. Two more rounds have been completed in October 2020 and February 2021. Each round participants were requested to return a self-collected finger-prick blood sample in a microtainer by mail and complete a questionnaire. Questions covered sociodemographic factors, clinical data (type and date of onset of symptoms), virological findings if applicable (SARS-CoV-2 RT-qPCR testing, and date and result of testing; February 2021 round only) and data on COVID-19 vaccination if applicable (brand product, number of vaccinations and their dates; February 2021 round only). The study was ethically approved by the Medical Research Ethics Committees United MEC-U and registered under trial number NL8473. All participants above the age of 12 gave written informed consent. Both parents or legal guardians of participating children below the age of 16 years also gave written informed consent for participation of the child.

### Laboratory methods

#### SARS-CoV-2 RT-qPCR testing in the household cohort

All available samples in the household cohort were tested for presence of SARS-CoV-2 as previously described [13, 17]. The results of the naso- and oropharyngeal swab, oral fluid and feces specimens at T1 or T2 were combined to one result: RT-qPCR negative (all negative) or RT-qPCR positive (any positive). Index cases were considered RT-qPCR positive even if they tested negative at T1 and T2 as they would have tested RT-qPCR positive with local health authorities prior to enrolment in the study.

#### Multiplex immunoassay for Immunoglobulin G detection in the household and national cohorts

Serum was separated from blood clot and stored at -20°C until analysis. Total IgG to Spike S1 and Nucleoprotein was measured with a MIA as previously described [4]. Median fluorescence intensity measurements were expressed as binding antibody units per milliliter (BAU/ml) using 5-parameter logistic interpolation of the International Standard for human anti-SARS-CoV-2 immunoglobulin (20/136 NIBSC standard) [18].

### Statistical analyses

All statistical analyses were performed in R version 4.0.2 [19]. Calculation of seropositivity thresholds and associated assay performance is detailed in the Supplementary Methods. Sensitivity of seropositivity to Nucleoprotein and Spike S1 in detecting a past RT-qPCR-confirmed SARS-CoV-2 infection was determined in 1) hospitalized COVID-19 patients (Intensive Care Unit or ward), 2) mild COVID-19 patients (i.e., with COVID-19-related symptoms but not hospitalized), and 3) individuals with an asymptomatic infection. Specificity was determined in those who tested RT-qPCR negative. COVID-19-related symptoms were classified as fever, coughing, shortness of breath, loss of taste or smell, sore throat, headache, pain while breathing, runny nose, muscle ache, diarrhoea, (extreme) tiredness and/or nausea.

Data from the national and household cohort were analyzed jointly. We did not include repeated samples from the same individuals. Unvaccinated participants who underwent SARS-CoV-2 confirmatory testing between two weeks and 6 months prior to serological sampling were included in the study. Exclusion criteria were incomplete symptom data, serological evidence of SARS-CoV-2 exposure prior to testing or other testing than RT-qPCR such as rapid antigen tests (Supplementary Figure 2). The time since onset of symptoms was used to determine sensitivity over time since infection. For asymptomatic participants in the national cohort, the time since RT-qPCR testing date was used. In the household cohort, the time since onset of symptoms or diagnosis date for the index case was used if the time since onset of symptoms was unknown or in asymptomatic participants. For reference, sera from 27 hospitalized COVID-19 cases between 14 days and 2 months after onset of symptoms were analysed: 7 patients in the household cohort (Supplementary Figure 2A), 10 patients from the Erasmus Medical Centre in Rotterdam (Medical Ethical Committee number METC 06/282) and 10 patients from the Dijklander hospital in Hoorn.

To describe IgG to Spike S1 and Nucleoprotein in a COVID-19 vaccinated study population, data from the two cohorts were also combined (Supplementary Figure 2). Participants who reported to have completed one or two doses of COVID-19 vaccination were included. As nearly all participants had received Pfizer/BioNTech, participants with other vaccine brands were excluded. Furthermore, participants with incomplete vaccination information, such as vaccination dates, were excluded. Past infection with SARS-CoV-2 was based on RT-qPCR confirmation in the household cohort and Spike S1 seroconversion in a previous study round or self-reported RT-qPCR testing where available in the national cohort.

Sensitivity and specificity estimates, and their 95% confidence intervals (CIs) were calculated applying Receiver Operating Characteristic (ROC) curves using the pROC package in R (version 1.16.2 [20]). CIs were computed with 2,000 stratified bootstrap replicates. The Wilcoxon-Mann-Whitney test was used to compare IgG measurements between vaccinated participants who did vs. those who did not experience a prior SARS-CoV-2 infection.

## Results

### SARS-CoV-2 RT-qPCR-tested study population

A total of 352 mild and asymptomatic participants had tested RT-qPCR positive for SARS-CoV-2 and 1,161 negative (Table 1). The majority was female (61% of the RT-qPCR positives and 61% of the RT-qPCR negatives) and in the age category 22-65 years old (71% of the RT-qPCR positives and 68% of the RT-qPCR negatives). Most of the RT-qPCR positives experienced mild COVID-19-related symptoms (90%) compared to 56% of the RT-qPCR negatives, while 10% of the RT-qPCR positives and 44% of the RT-qPCR negatives were asymptomatic.

**Table 1:** General description of the RT-qPCR-confirmed SARS-CoV-2 study population.

| % (n) | SARS-CoV-2 positive | SARS-CoV-2 negative |
| --- | --- | --- |
| N* | 352 | 1,161 |
| Female | 61.1% (215) | 60.5% (702) |
| Age category (years) |  |  |
| - 1-21 | 17.1% (60) | 16.0% (186) |
| - 22-65 | 71.0% (250) | 68.4% (794) |
| - 66-87 | 11.9% (42) | 15.6% (181) |
| COVID-19 related symptoms** |  |  |
| - No | 10.2% (36) | 43.6% (506) |
| - Yes | 89.8% (316) | 56.4% (655) |

\*See Supplementary Figure 2A and 2B for more information on sample availability and exclusion criteria. \*\*Fever, coughing, shortness of breath, loss of taste or smell, sore throat, headache, pain while breathing, runny nose, muscle ache, diarrhoea, (extreme) tiredness and/or nausea.

### Nucleoprotein and Spike S1 seropositivity to detect past SARS-CoV-2 infection

Nucleoprotein and Spike S1 IgG measurements by SARS-CoV-2 RT-qPCR status and symptom status are shown in Figure 1A. Sensitivity of Nucleoprotein was highest in hospitalized COVID-19 patients (100%) between two weeks and two months post onset of symptoms as compared to mild COVID-19 (79%, 95% CI: 75-84%) or asymptomatic SARS-CoV-2 (67%, 50-81%) between two weeks and six months following symptom onset/infection (Figure 1B). Seropositivity to Spike S1 showed higher sensitivity estimates (i.e., hospitalized: 100%, mild COVID-19: 89%, 85-92%, asymptomatic SARS-CoV-2: 72%, 56-86%). Sensitivity of Nucleoprotein for mild COVID-19 was highest shortly after infection: 85% (79-91%) at 2 weeks to 2 months following the onset of symptoms to 79% (70-86%) at 3-4 months and 59% (44-72%) at 5-6 months. This decline was faster than that seen for Spike S1 (from 90%, 85-94%, to 90%, 83-95% to 80%, 67-91%; Figure 1C).

**Figure 1:**
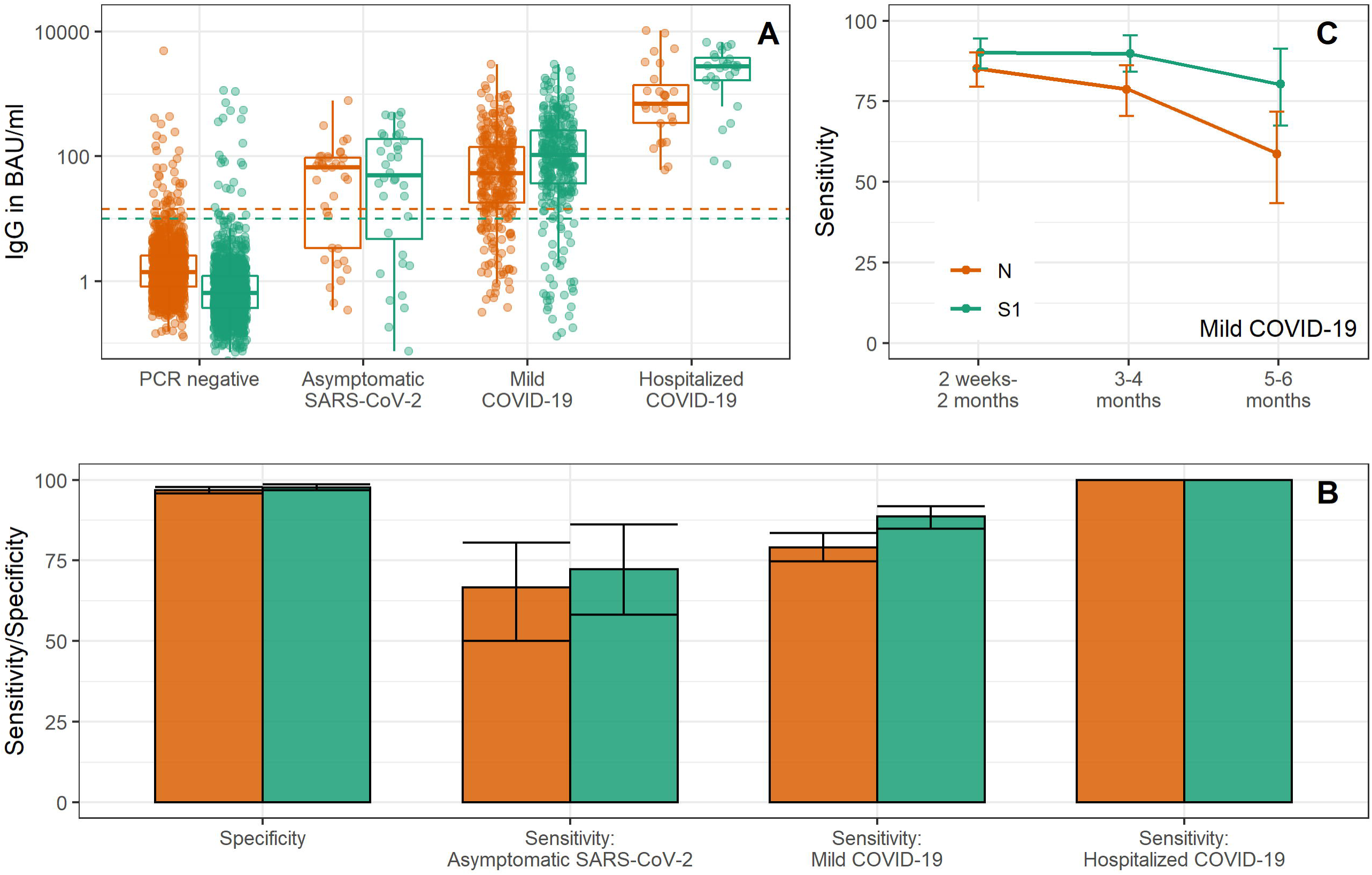
Nucleoprotein and Spike S1 IgG responses to detect SARS-CoV-2 infections. In (A) IgG measurements to Nucleoprotein and Spike S1 are shown by SARS-CoV-2 PCR status and symptom status along with the threshold for seropositivity (dashed horizontal line). In (B) specificity and sensitivity estimates with 95% confidence intervals are shown for Nucleoprotein and Spike S1 seropositivity. In (C) sensitivity estimates with 95% confidence intervals of Nucleoprotein and Spike S1 seropositivity over time (in months); this does not include repeated samples from the same individuals. S1: Spike S1, N: Nucleoprotein, IgG: immunoglobulin G, BAU/ml: binding antibody units.

Specificity in RT-qPCR negative tested persons was 97% (96-98%) for Nucleoprotein and 98% (97-99%) for Spike S1 (Figure 1B). For persons who seroconverted to either Nucleoprotein or Spike S1 within the RT-qPCR-negative selection (n=46), 19 had seroconverted to Nucleoprotein only (41%), 9 to Spike S1 only (20%) and 18 to both Nucleoprotein and Spike S1 (39%).

### Spike S1 and Nucleoprotein IgG kinetics after COVID-19 vaccination with Pfizer/BioNTech

Of the 230 Pfizer/BioNTech vaccinated participants, 118 had received two doses at the time of sampling (51%), 172 were female (75%) and 177 were 18-65 years old (77%) vs. 53 who were >65 years old (23%) (Table 2). In previously naïve individuals (n=179), IgG to Spike S1 showed a homogenous response between individuals over time since vaccination (Figure 2A). After two weeks, 96% of the previously naïve individuals had seroconverted to Spike S1 (126/131, Figure 2A). The majority was seronegative for Nucleoprotein (93%, 122/131; Figure 2B). Of the nine seropositive individuals, four were already seropositive for Nucleoprotein prior to vaccination but not for Spike S1. Participants who had experienced a SARS-CoV-2 infection preceding vaccination, produced 2.7-fold higher median levels of IgG to Spike S1 ≥14 days after the first dose as compared to those unexposed to SARS-CoV-2 at ≥7 days after the second dose (6,480 vs. 2,438 BAU/ml, p=0.011, Figure 2C).

**Figure 2:**
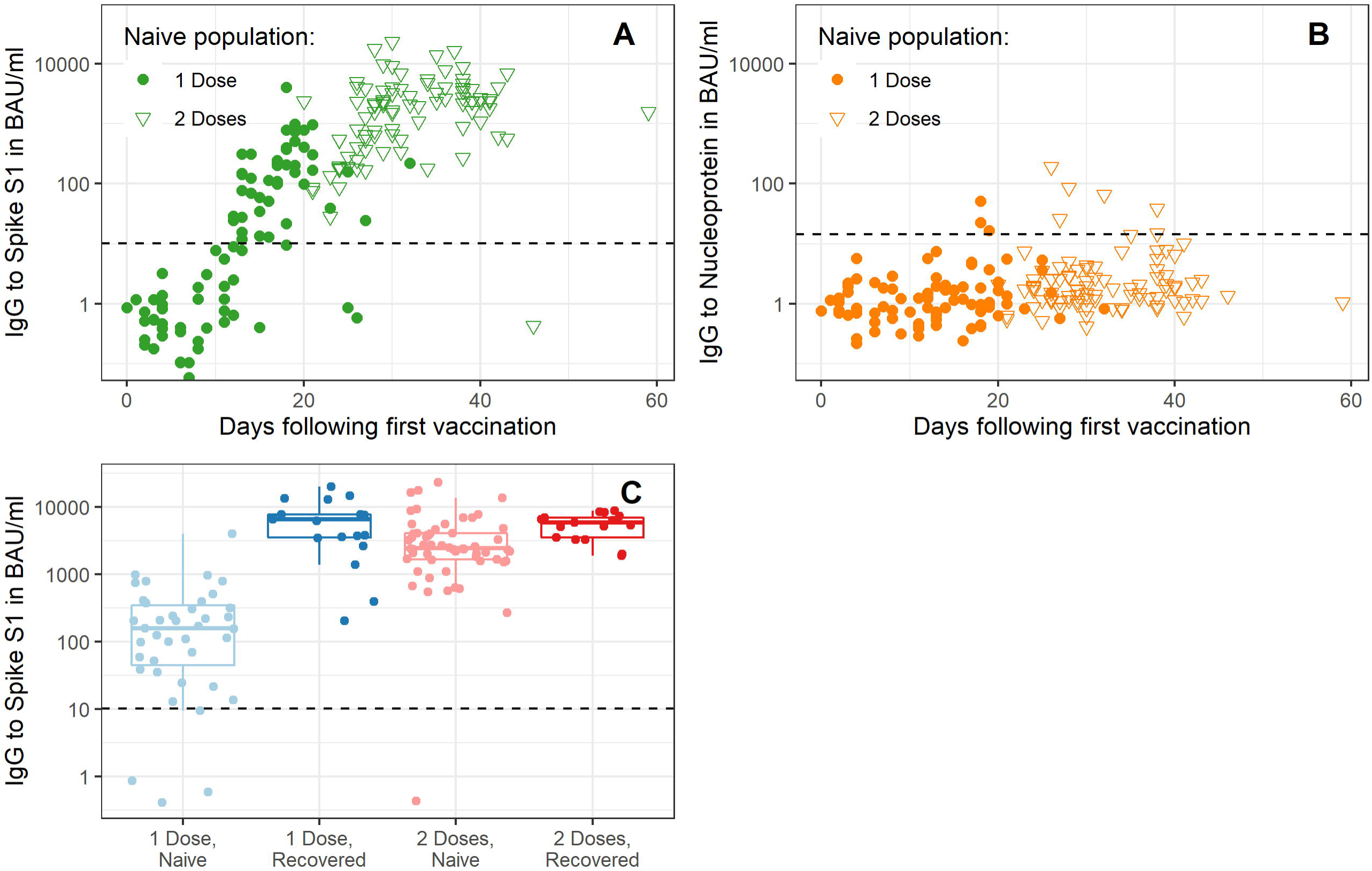
Nucleoprotein and Spike S1 IgG kinetics following COVID-19 vaccination. IgG measurements to Spike S1 (A) and Nucleoprotein (B) in naïve individuals are shown over days since first vaccination. In (C) IgG measurements to Spike S1 are shown by prior exposure status and number of doses received, individuals were included if they were sampled ≥14 days after the first dose or ≥7 days after the second dose. In (A-C) the dashed horizontal line depicts the threshold for seropositivity. S1: Spike S1, N: Nucleoprotein, IgG: immunoglobulin G, BAU/ml: binding antibody units.

**Table 2:** General description of the COVID-19 vaccinated study population.

|  | % (n)<br>unless otherwise specified |
| --- | --- |
| N* | 230 |
| Female | 75% (172) |
| Age category in years |  |
| - 18-65 | 77 (177) |
| - 66-91 | 23% (53) |
| Healthcare worker | 75% (172) |
| Vaccination |  |
| - Two doses | 51% (118) |
| - Median days since first vaccination, range (IQR) | 24, 0-59 (14-32) |
| Past SARS-CoV-2 infection |  |
| - No | 78% (179) |
| - (Self-reported) RT-qPCR positive | 14% (32) |
| - Seroconversion IgG to Spike S1 prior to vaccination | 8% (19) |

\*See Supplementary Figure 2A and 2B for more information on sample availability and exclusion criteria. \*\*Fever, coughing, shortness of breath, loss of taste or smell, sore throat, headache, pain while breathing, runny nose, muscle ache, diarrhoea, (extreme) tiredness and/or nausea.
IQR: interquartile range; IgG: immunoglobulin G.

Of the COVID-19 vaccinated participants with evidence for a previous SARS-CoV-2 infection, 13 experienced symptom onset less than five months prior to sampling. Ten of these 13 had seroconverted to Nucleoprotein (77%, 54-100%), while nearly all were symptomatic during their infection (92%, 12/13). At 8-11 months post symptom onset, 45% (29-61%) were seropositive for Nucleoprotein while 82% (31/38) had been symptomatic during their infection.

## Discussion

As COVID-19 vaccines induce immune responses to the Spike protein, alternative serological targets need to be considered for serosurveillance of SARS-CoV-2 breakthrough infections. Here, we showed that seropositivity to Nucleoprotein can detect mild COVID-19 with a sensitivity of 85% between two weeks and two months following symptom onset compared to 90% for Spike S1. At 3-4 months post symptom onset, sensitivity declined to 79% for Nucleoprotein while it remained 90% for Spike S1.

Several publications have focused on the sensitivity of Nucleoprotein to detect past RT-qPCR-confirmed SARS-CoV-2 infections with estimates ranging from 70% to 96% [6-9]. The wide range in the observed sensitivity estimates is likely due to differences in the used reference population, often consisting of hospitalized patients or healthcare workers with COVID-19, and differences related to the applied antibody detection platforms. Few have stratified results by symptomatic status of the reference populations or time since onset of symptoms [7]. This while breakthrough infections are expected to be even more frequently mild or asymptomatic than primary infections. In mildly symptomatic persons (healthcare workers), Mariën et al. reported a sensitivity of 70% within six weeks and 85% more than five months after symptom onset [7]. We also reported a sensitivity of 85% in mildly symptomatic patients between two weeks and two months after symptom onset though we saw a decline to 59% at 5-6 months after symptom onset. In persons with an asymptomatic SARS-CoV-2 infection in our study, sensitivity of Nucleoprotein was 67%; insufficient numbers were available to stratify this estimate further by time since infection. Our estimate for Spike S1 sensitivity in asymptomatic participants (72%; 26/36) was similar to that recently published by Vanshylla et al. combining IgG and IgA responses (77%; 34/44) [21]. Although Nucleoprotein sensitivity was lower in asymptomatic individuals, few to none of these individuals would have been identified by passive surveillance using RT-qPCR or antigen tests as the absence of symptoms would limit chances of seeking diagnosis.

Sensitivity decreased to 79% at 3-4 months following infection, a 8% seroreversion in 2 months. Choudhry et al. reported 31% seroreversion for Nucleoprotein IgG after three months in seroconverted healthcare workers in the United Kingdom using rapid IgG/IgM tests [22]. Others have likewise shown that Nucleoprotein IgG antibodies decline on average 1.5-2 times faster than those to Spike S1 [10, 23]. The higher rate of seroreversion for Nucleoprotein compared to Spike S1, means that regular serological measurements is recommended to ensure detection of breakthrough infections (e.g. 2-3 monthly intervals). The serological response to Nucleoprotein may vary more following COVID-19 vaccination as partial immunity might limit viral replication and thus exposure of the immune system to the viral Nucleoprotein. Asymptomatic and mild cases that might be missed by RT-qPCR or serological testing are unlikely to pose a risk of development of disease requiring hospitalization. However, they might still contribute to transmission of the virus.

Previous specificity estimates for bead-based assays were ≥97% based on pre-pandemic controls [7-9]. Here we likewise showed that 97% of the RT-qPCR-negative population was seronegative for Nucleoprotein. However, presence of SARS-CoV-2 could have been missed in our study due to no detectable SARS-CoV-2 RNA at the time of sampling or incorrect sampling thus lowering specificity. 18 out of 37 Nucleoprotein seropositive persons also seroconverted to Spike S1 which strengthens the hypothesis that these samples represent participants not being sampled optimally for RT-qPCR. The performance of an assay is a trade-off between sensitivity and specificity. As we expect the prevalence of breakthrough infections to be low, we focused on high specificity by setting a conservative seropositivity threshold.

Most of the COVID-19 vaccinated participants in the current study were healthcare workers who received Pfizer/BioNTech. IgG to Spike S1 after two doses showed comparable levels as those for healthcare workers from another study conducted in Rotterdam, the Netherlands [24], while a previously exposed population already produced robust IgG after one dose of Pfizer/BioNTech vaccine. Interestingly, the majority of the previously infected participants receiving one vaccination dose got infected approximately 11 months prior to vaccination (72%, 18/25). This suggests that infection up to a year prior to vaccination still enables robust boosting of IgG to Spike S1 as observed by others [25, 26], though numbers were small and nearly all were symptomatic. We confirmed that Nucleoprotein also detects past SARS-CoV-2 infection in the previous five months in this vaccinated population (77%), though the confidence interval was wide due to the low numbers available.

There are strengths and weaknesses in the cohorts we used in this study. Study team nurses collecting samples and questionnaire data at pre-set sampling timepoints is the strength of the household cohort, but its weakness includes that results are likely to correlate within families (i.e., genetic relatedness and immune response). The national cohort is more representative of the general population, including more asymptomatic individuals, and the repeated cross-sectional design ensured that participants were included with different time frames since infection and/or vaccination. However, the weakness of this approach is that it relied on self-reported data. Several types of bias may arise from self-reported data including recall bias, e.g. those who tested SARS-CoV-2 positive might be more likely to remember the type of symptoms or test they received.

In conclusion, we showed that Nucleoprotein can detect prior SARS-CoV-2 infections with a sensitivity of 85% in a mildly symptomatic unvaccinated population between two weeks and two months after symptom onset. Serological responses to Nucleoprotein may thus prove helpful in identifying the frequency of SARS-CoV-2 infections in vaccinated persons, alongside molecular tests. Furthermore, it can help to interpret IgG to Spike S1 responses after COVID-19 vaccination as particularly high responses shortly after vaccination could be explained by prior exposure history.

## Data Availability

Data are available from the authors upon reasonable request.

## Acknowledgments

We would like to thank all study participants and the team at the laboratory conducting all RT-qPCR assays, represented by Bas van der Veer, Sharon van den Brink and Anne-Marie van den Brandt.

## References

1. Dong, E., H. Du, and L. Gardner, An interactive web-based dashboard to track COVID-19 in real time. Lancet Infect Dis, 2020. 20(5): p. 533–534.

2. Polack, F.P., et al., Safety and Efficacy of the BNT162b2 mRNA Covid-19 Vaccine. N Engl J Med, 2020. 383(27): p. 2603–2615.

3. Sheikh, A., et al., SARS-CoV-2 Delta VOC in Scotland: demographics, risk of hospital admission, and vaccine effectiveness. Lancet, 2021. 397(10293): p. 2461–2462.

4. den Hartog, G., et al., SARS-CoV-2-Specific Antibody Detection for Seroepidemiology: A Multiplex Analysis Approach Accounting for Accurate Seroprevalence. J Infect Dis, 2020. 222(9): p. 1452–1461.

5. den Hartog, G., et al., Persistence of antibodies to SARS-CoV-2 in relation to symptoms in a nationwide prospective study. Clin Infect Dis, 2021.

6. Fenwick, C., et al., Changes in SARS-CoV-2 Spike versus Nucleoprotein Antibody Responses Impact the Estimates of Infections in Population-Based Seroprevalence Studies. J Virol, 2021. 95(3).

7. Marien, J., et al., Evaluating SARS-CoV-2 spike and nucleocapsid proteins as targets for antibody detection in severe and mild COVID-19 cases using a Luminex bead-based assay. J Virol Methods, 2021. 288: p. 114025.

8. Rosado, J., et al., Multiplex assays for the identification of serological signatures of SARS-CoV-2 infection: an antibody-based diagnostic and machine learning study. Lancet Microbe, 2021. 2(2): p. e60–e69.

9. Fotis, C., et al., Accurate SARS-CoV-2 seroprevalence surveys require robust multi-antigen assays. Sci Rep, 2021. 11(1): p. 6614.

10. Dan, J.M., et al., Immunological memory to SARS-CoV-2 assessed for up to 8 months after infection. Science, 2021. 371(6529).

11. Wu, Z. and J.M. McGoogan, Characteristics of and Important Lessons From the Coronavirus Disease 2019 (COVID-19) Outbreak in China: Summary of a Report of 72314 Cases From the Chinese Center for Disease Control and Prevention. JAMA, 2020. 323(13): p. 1239–1242.

12. McDonald, S.A., et al., Estimating the asymptomatic proportion of SARS-CoV-2 infection in the general population: Analysis of nationwide serosurvey data in the Netherlands. Eur J Epidemiol, 2021.

13. Reukers, D.F.M., et al., High infection secondary attack rates of SARS-CoV-2 in Dutch households revealed by dense sampling. Clin Infect Dis, 2021.

14. Vos, E.R.A., et al., Nationwide seroprevalence of SARS-CoV-2 and identification of risk factors in the general population of the Netherlands during the first epidemic wave. J Epidemiol Community Health, 2020.

15. Vos, E.R.A., et al., Associations between measures of social distancing and SARS-CoV-2 seropositivity: a nationwide population-based study in the Netherlands. Clin Infect Dis, 2021.

16. Verberk, J.D.M., et al., Third national biobank for population-based seroprevalence studies in the Netherlands, including the Caribbean Netherlands. BMC Infect Dis, 2019. 19(1): p. 470.

17. Corman, V.M., et al., Detection of 2019 novel coronavirus (2019-nCoV) by real-time RT-PCR. Euro Surveill, 2020. 25(3).

18. World Health Organization. First WHO International Standard for anti-SARS-CoV-2 immunoglobulin (human). 2020 [cited 2021 7 June 2021]; Available from: https://www.nibsc.org/documents/ifu/20-136.pdf.

19. R Core Team, R: A language and environment for statistical computing. 2020, R Foundation for Statistical Computing: Vienna, Austria.

20. Robin, X., et al., pROC: an open-source package for R and S+ to analyze and compare ROC curves. BMC Bioinformatics, 2011. 12: p. 77.

21. Vanshylla, K., et al., Kinetics and correlates of the neutralizing antibody response to SARS-CoV-2 infection in humans. Cell Host Microbe, 2021.

22. Choudhry, N., et al., Disparities of SARS-CoV-2 Nucleoprotein-Specific IgG in Healthcare Workers in East London, UK. Front Med (Lausanne), 2021. 8: p. 642723.

23. Wheatley, A.K., et al., Evolution of immune responses to SARS-CoV-2 in mild-moderate COVID-19. Nat Commun, 2021. 12(1): p. 1162.

24. Geers, D., et al., SARS-CoV-2 variants of concern partially escape humoral but not T-cell responses in COVID-19 convalescent donors and vaccinees. Sci Immunol, 2021. 6(59).

25. Favresse, J., et al., Early antibody response in health-care professionals after two doses of SARS-CoV-2 mRNA vaccine (BNT162b2). Clin Microbiol Infect, 2021.

26. Abu Jabal, K., et al., Impact of age, ethnicity, sex and prior infection status on immunogenicity following a single dose of the BNT162b2 mRNA COVID-19 vaccine: real-world evidence from healthcare workers, Israel, December 2020 to January 2021. Euro Surveill, 2021. 26(6).

