## Supplementary Methods for "Seropositivity to Nucleoprotein to detect SARS-CoV-2 infections: a tool to detect breakthrough infections after COVID-19 vaccination"

Threshold for seropositivity and assay performance…………………………..…………………………………… page 2
Supplementary Figure 1: Density distribution of log-transformed immunoglobulin G (IgG) measurements…………………………………………………………………………..……………………………………………. page 3

Supplementary Figure 2: Flowchart of sample availability and exclusion criteria for the household and national cohorts………………………………………………………………………………………………………………… page 4

References………………………………………………………………………………………………………………………………. page 7

***Threshold for seropositivity and assay performance***

We have previously reported the clinical performance of the MIA assay using a reference panel of laboratory-confirmed COVID-19 cases (38% hospitalized), pre-pandemic negative controls and cases with laboratory-confirmed respiratory infections other than SARS-CoV-2, including the four seasonal human coronaviruses. We found a 99.0% specificity and 84.4% sensitivity for Spike S1 IgG, and 98.5% specificity and 89.4% sensitivity for Nucleoprotein IgG [1]. Since then, we optimized the assay for Spike S1 and the seropositivity threshold for Spike S1 by applying a mixture model to IgG measurements from the June 2020 round of the national survey aiming for higher specificity [2, 3]. This led to 99.1% specificity and 91.6% sensitivity for Spike S1 using the aforementioned reference panel [2].

The seropositivity threshold for Nucleoprotein was also defined by applying a mixture model to the natural log-transformed IgG measurements in the June 2020 study round of the national cohort using the *mixtools* package (version 1.2.0 [4]). Normal distributions were assumed for the seronegative population (lower distribution, mean: 0.587; standard deviation: 0.807) and the seropositive population (upper distribution, mean: 3.041; standard deviation: 1.839).

The threshold for seropositivity was set at the 99.9^th^ percentile of the lower normal distribution. This resulted in the following seropositivity thresholds (on the linear scale): 10.08 BAU/ml for Spike S1 [3], note that this is the same threshold as previously reported at 1.04 which has now been updated to BAU/ml, and 14.29 BAU/ml for Nucleoprotein (Supplementary Figure 1).

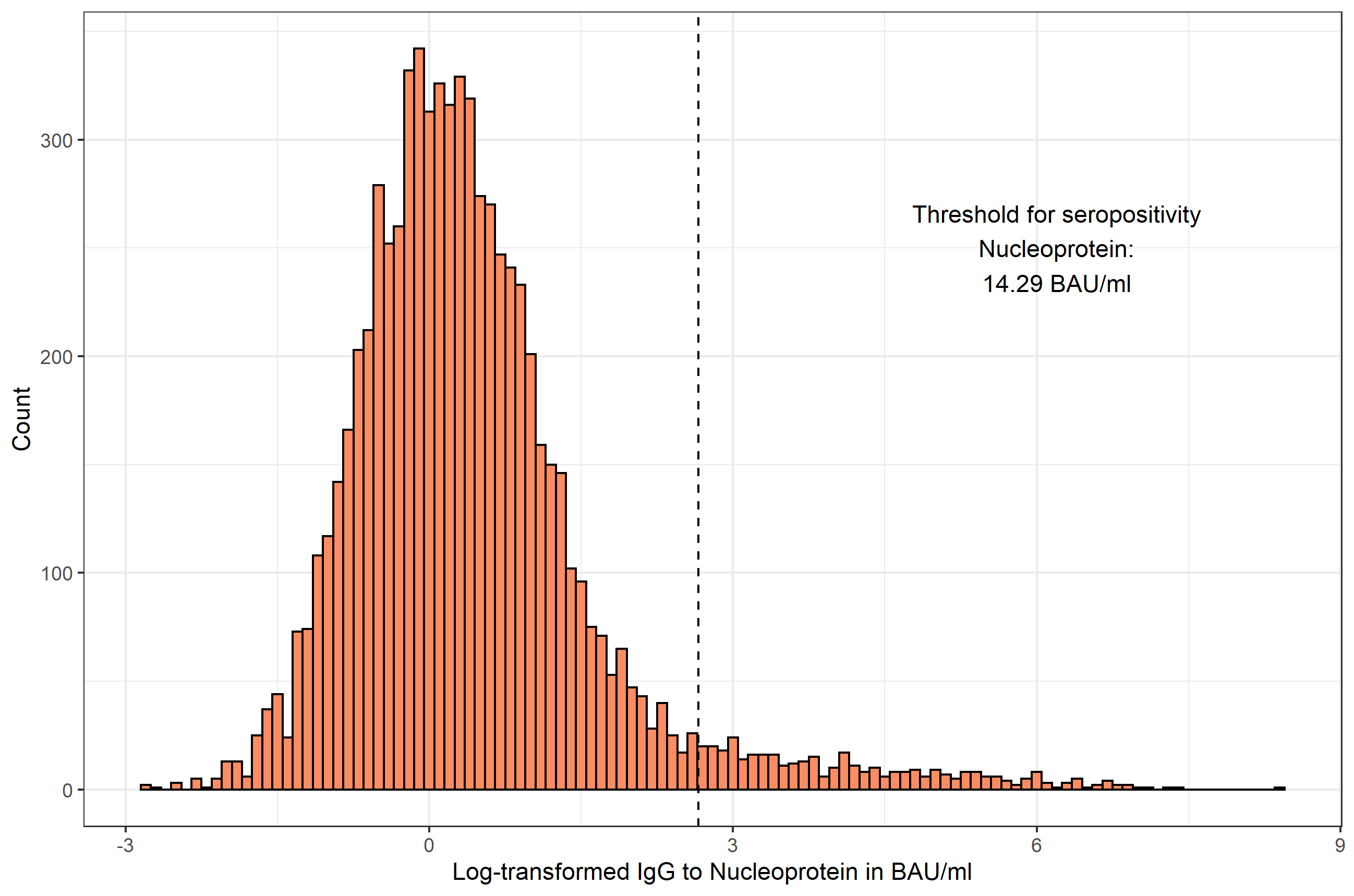

**Supplementary Figure 1: Density distribution of log-transformed immunoglobulin G (IgG) measurements.** IgG is expressed as natural log-transformed binding arbitrary units per ml (BAU/ml). The threshold for seropositivity is shown as the vertical dashed line.

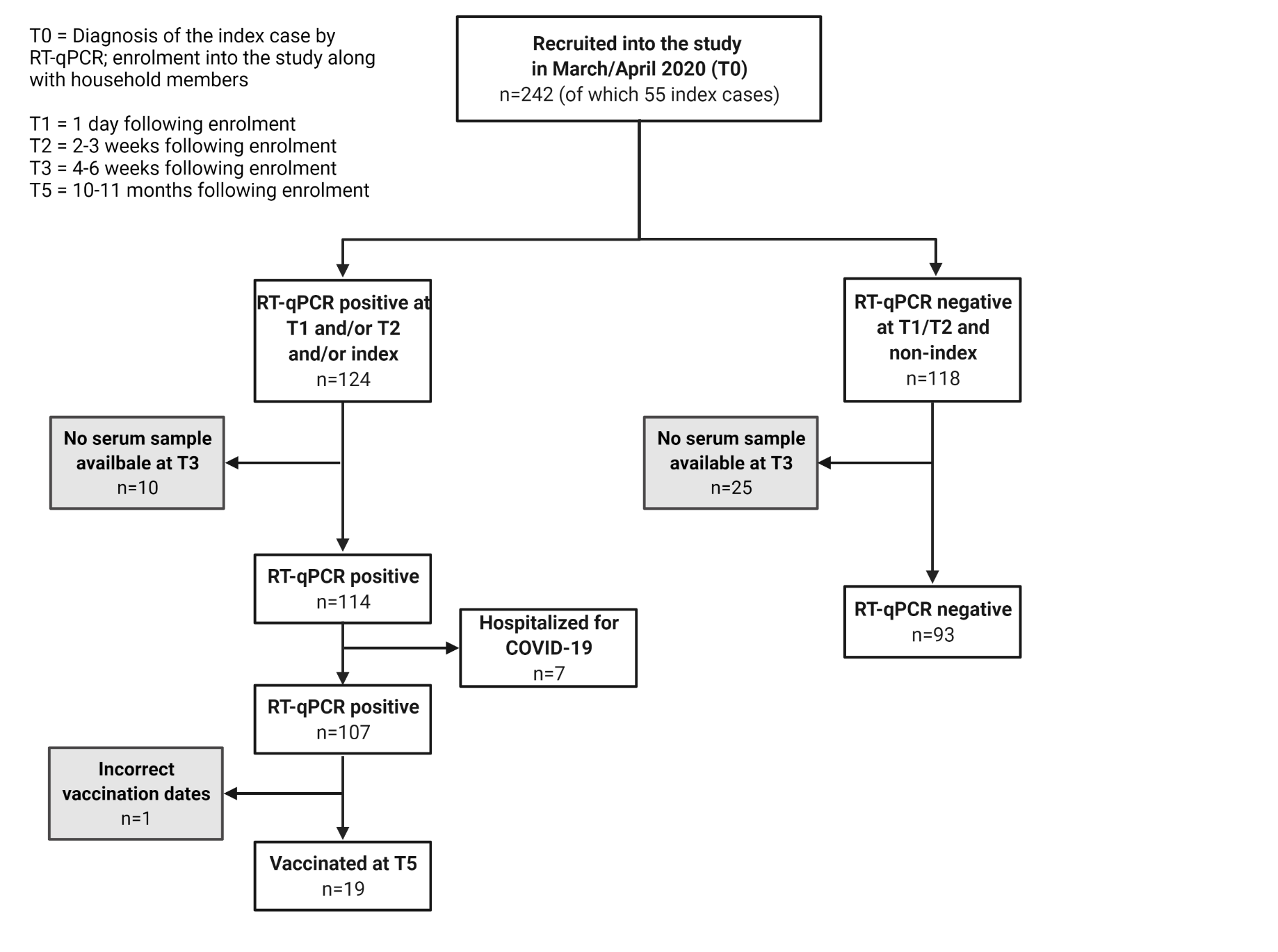

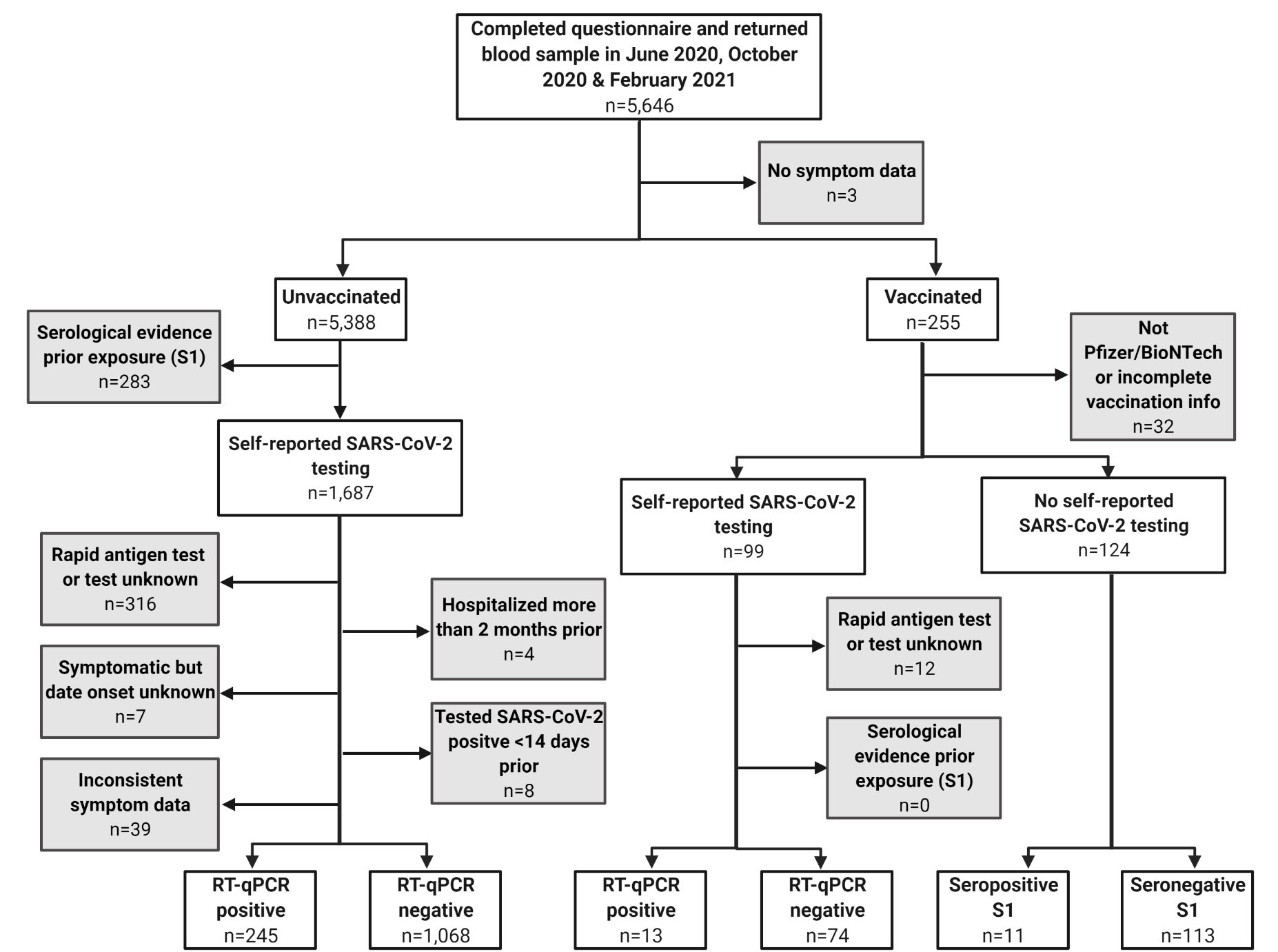

**Supplementary Figure 2: Flowchart of sample availability and exclusion criteria for the household (A) and national (B) cohorts.** Excluded participants are shown in gray boxes. RT-qPCR: reverse transcription quantitative polymerase chain reaction (i.e., molecular test for presence of SARS-CoV-2 viral RNA); S1: Spike S1.

**References**

1.	den Hartog, G., et al., *SARS-CoV-2-Specific Antibody Detection for Seroepidemiology: A Multiplex Analysis Approach Accounting for Accurate Seroprevalence.* J Infect Dis, 2020. **222**(9): p. 1452-1461.

2.	den Hartog, G., et al., *Persistence of antibodies to SARS-CoV-2 in relation to symptoms in a nationwide prospective study.* Clin Infect Dis, 2021.

3.	Vos, E.R.A., et al., *Associations between measures of social distancing and SARS-CoV-2 seropositivity: a nationwide population-based study in the Netherlands.* Clin Infect Dis, 2021.

4.	Benaglia, T., et al., *mixtools: An R Package for Analyzing Mixture Models.* Journal of Statistical Software; Vol 1, Issue 6 (2010), 2009.

5.	Hou, H., et al., *Detection of IgM and IgG antibodies in patients with coronavirus disease 2019.* Clin Transl Immunology, 2020. **9**(5): p. e01136.
